# Detecting CTP Truncation Artifacts in Acute Stroke Imaging from the Arterial Input and the Vascular Output Functions

**DOI:** 10.1101/2022.06.16.22276371

**Authors:** Ezequiel de la Rosa, Diana M. Sima, Jan S. Kirschke, Bjoern Menze, David Robben

## Abstract

**Background:** Current guidelines for CT perfusion (CTP) in acute stroke suggest acquiring scans with a minimal duration of 60-70 s. But even then, CTP analysis can be affected by truncation artifacts. Conversely, shorter acquisitions are still widely used in clinical practice and are usually sufficient to reliably estimate lesion volumes. We aim to devise an automatic method that detects scans affected by truncation artifacts.

**Methods:** Shorter scan durations are simulated from the ISLES’18 dataset by consecutively removing the last CTP time-point until reaching a 10 s duration. For each truncated series, perfusion lesion volumes are quantified and used to label the series as *unreliable* if the lesion volumes considerably deviate from the original untruncated ones. Afterwards, nine features from the arterial input function (AIF) and the vascular output function (VOF) are derived and used to fit machine-learning models with the goal of detecting unreliably truncated scans. Methods are compared against a baseline classifier solely based on the scan duration, which is the current clinical standard. The ROC-AUC, precision-recall AUC and the F1-score are measured in a 5-fold cross-validation setting.

**Results:** Machine learning models obtained high performance, with a ROC-AUC of 0.964 and precision-recall AUC of 0.958 for the best performing classifier. The highest detection rate is obtained with support vector machines (F1-score = 0.913). The most important feature is the AIF_coverage_, measured as the time difference between the scan duration and the AIF peak. In comparison, the baseline classifier yielded a lower performance of 0.940 ROC-AUC and 0.933 precision-recall AUC. At the 60-second cutoff, the baseline classifier obtained a low detection of unreliably truncated scans (F1-Score = 0.638).

**Conclusions:** Machine learning models fed with discriminant AIF and VOF features accurately detected unreliable stroke lesion measurements due to insufficient acquisition duration. Unlike the 60s scan duration criterion, the devised models are robust to variable contrast injection and CTP acquisition protocols and could hence be used for quality assurance in CTP post-processing software.

## Introduction

Treatment decision making in acute ischemic stroke is mostly guided by computed tomography (CT) imaging, as the technique allows to answer (at least) four crucial questions regarding the patient brain’s condition: 1) Is there hemorrhage? 2) Is there any thrombus that could be targeted? 3) Is there already irreversibly damaged tissue (a.k.a. *core*)? 4) Is there salvageable tissue (a.k.a. *penumbra*, tissue at risk but potentially recoverable)? Konstas et al. (2009). While the first two questions can be answered with non-contrast CT and CT angiography, respectively, the last two questions are typically addressed through CT perfusion (CTP). CTP is of major importance for neuroradiologists as it allows the identification of patients that could benefit from recanalization therapies Albers et al. (2016). In this context, distinguishing potentially salvageable brain tissue from already necrosed areas drive the therapheutical decision making.

In clinical routine, CTP post-processing software is used to estimate perfusion maps and to quantify perfusion lesion volumes. The perfusion maps used in acute ischemic stroke are derived from the CTP contrast attenuation curves and are cerebral blood volume, cerebral blood flow (CBF), mean transit time and time to the maximum of the residue function (Tmax). There exist several different techniques implemented in clinical and/or research software packages to estimate these perfusion metrics. Among the most widely used are the Fourier transform and the delay-invariant singular value decomposition deconvolution techniques using time-shift Smith et al. (2004) or block-circulant approaches Wu et al. (2003); Wittsack et al. (2008). Independently of their functioning, the end goal of CTP software packages is the accurate quantification of perfusion maps and, consequently, the reliable volumetric quantification of the brain lesions. Despite the vast adoption of CT perfusion software in clinical routine, there are well known and persistent pitfalls of these techniques that hamper the brain lesion quantification and hence their interpretation, as described in Mangla et al. (2014); Potter et al. (2019); Vagal et al. (2019); Chung et al. (2021). This work focuses on the so called *truncation* of the time attenuation curves, which could be defined as the early ending of the CTP acquisition that precludes the entire capture of the tissue perfusion phases Vagal et al. (2019).

CTP truncation artifacts have extensively been observed in previous works Campbell et al. (2011); Kamalian et al. (2012); d’Esterre et al. (2015); Mikkelsen et al. (2015); Geuskens et al. (2015); Borst et al. (2015); Copen et al. (2015); Kasasbeh et al. (2016). As described in practical acute stroke imaging recommendations, the CTP analysis should include a quality control step that checks for complete acquisition of the perfusion curves including both the contrast agent wash-in and wash-out phases Vagal et al. (2019); Christensen and Lansberg (2019); Chung et al. (2021). Visual identification of truncated AIF/VOF and/or time attenuation curves has been conducted in previous studies Geuskens et al. (2015); Borst et al. (2015). Despite the fact that visual quality control could easily detect truncated perfusion curves, it is not straight-forward to understand the implications of such curves truncations over the quantified lesion volumes. Thus, finding whether the truncation effects are strong enough to considerably perturb the quantified perfusion volumes could only be assessed through quantitative analyses. A major step in understanding the quantitative impact of truncation artifacts over the perfusion maps was done in Copen et al. (2015). The work showed that truncation artifacts depend on the truncation degree and affect the perfusion metrics differently depending on the used deconvolution algorithm. Moreover, the CTP truncation effects over the brain lesion volumes were studied in Kasasbeh et al. (2016). The authors found that a 60 second scan duration is enough to avoid volumetric errors in 95% of their analysed scans. These results have later been adopted as a practical recommendation for the implementation of CTP in acute stroke Christensen and Lansberg (2019). In clinical routine, however, different centers or scanner operators make use of post-processing software from different vendors (and with diverse deconvolution algorithms), as well as different contrast injection and CTP acquisition protocols. Shorter acquisitions are frequently adopted by centers in order to reduce the exposure of the patient to ionizing radiation under the ALARA (i.e. as low as reasonably achievable) principle. Based on these considerations, it is possible that scans with shorter than 60 second scan duration could reliably estimate lesion volumes while scans with different characteristics could suffer from truncation errors even while having a 60-70 second acquisition duration.

In this work we propose a tool for the automatic identification of unreliable perfusion volumes due to insufficient scan duration. Our proposal makes use of simple and easy to extract features derived from the vascular perfusion curves (i.e. the arterial input function, AIF, and the vascular output function, VOF). Experiments on the public ISLES’18 dataset show that truncation artifacts impact the perfusion-derived features, hence allowing their identification with machine learning models. The proposed approach increases the interpretability of acute ischemic stroke outputs obtained in clinical practice with CTP post-processing software.

## Materials and methods

### Data

The ISLES’18 dataset is used for our experiments Cereda et al. (2016); Hakim et al. (2021). The database is multi-center and multi-scanner and includes 156 CTP scans obtained from 103 acute stroke patients. For our experiments, we have used the preprocessed scans from the ISLES 2018 challenge (http://www.isles-challenge.org/). The CTP volumes have been motion corrected, coregistered and spatio-temporally resampled (256 *×* 256 matrix, 1 volume per second). A full dataset description can be found in Cereda et al. (2016).

### Simulating Shorter CTP Scans

We simulate shorter CTP scan durations by repeatedly discarding a 1 second timepoint from the end of the series until reaching the 10 first seconds of it. Note that the number of truncated simulated series varies from scan to scan, depending on its original total duration.

### CTP Post-processing

Each truncated CTP series is analyzed using a research version of ico**brain cva** 1.4.1 (icometrix, Leuven, Belgium), an FDA-cleared and CE-marked software for acute stroke CTP post-processing. Each truncated series is processed using experts’ manually annotated vascular functions available in de la Rosa et al. (2021). Please note that the manual AIF/VOF does not change location for all shorter versions of a same scan. The vascular functions from each truncated scan are retained for the subsequent experiments.

Perfusion maps (Tmax, CBF, cerebral blood volume and mean transit time) are obtained through delay-invariant singular value decomposition deconvolution. Absolute and relative CBF maps are computed, where the relative rCBF map is obtained after normalization of the absolute one using mean control tissue values. Control tissue is defined by the software as Tmax *<* 6s Lin et al. (2016). Quantification of the hypoperfused and core lesion volumes is automatically obtained by the software using Tmax *>* 6s Lin et al. (2016) and rCBF *<* 0.38 (within the hypoperfused tissue area), respectively. The used rCBF cutoff (which is set in the software just for the purpose of these experiments) has been identified as optimal for the ISLES’18 dataset Cereda et al. (2016).

### Defining Truncation Artifacts

In order to label each shorter scan version as *reliable* or *unreliable* (i.e., considerably suffering from truncation artifacts), we first check that the original unshortened scan does not already suffer from truncation artifacts. As such, scans are labeled to be *stable* if truncation of the final 6 frames or less did not impact the computed volumes by more than 2.5 ml Kasasbeh et al. (2016); otherwise, scans are labelled as *unstable* ones. For our experiments, all unstable scans have been discarded from further analyses. Besides, scans without a hypoperfused lesion have also been excluded as their stability can not be guaranteed.

The truncated series from all stable scans are labelled as *reliable* if the corresponding hypoperfused and core volumes deviated *<*10% or *<* 5 ml from the untruncated volume estimates. Otherwise, the truncated scan (and all its shorter versions) are labelled as *unreliable*. Besides, for each CTP series, the optimal scan duration is defined as the shortest scan duration providing reliable volumes estimates. Figure 1 shows a stable CTP scan example with its corresponding reliability truncation labels.

**Figure 1:**
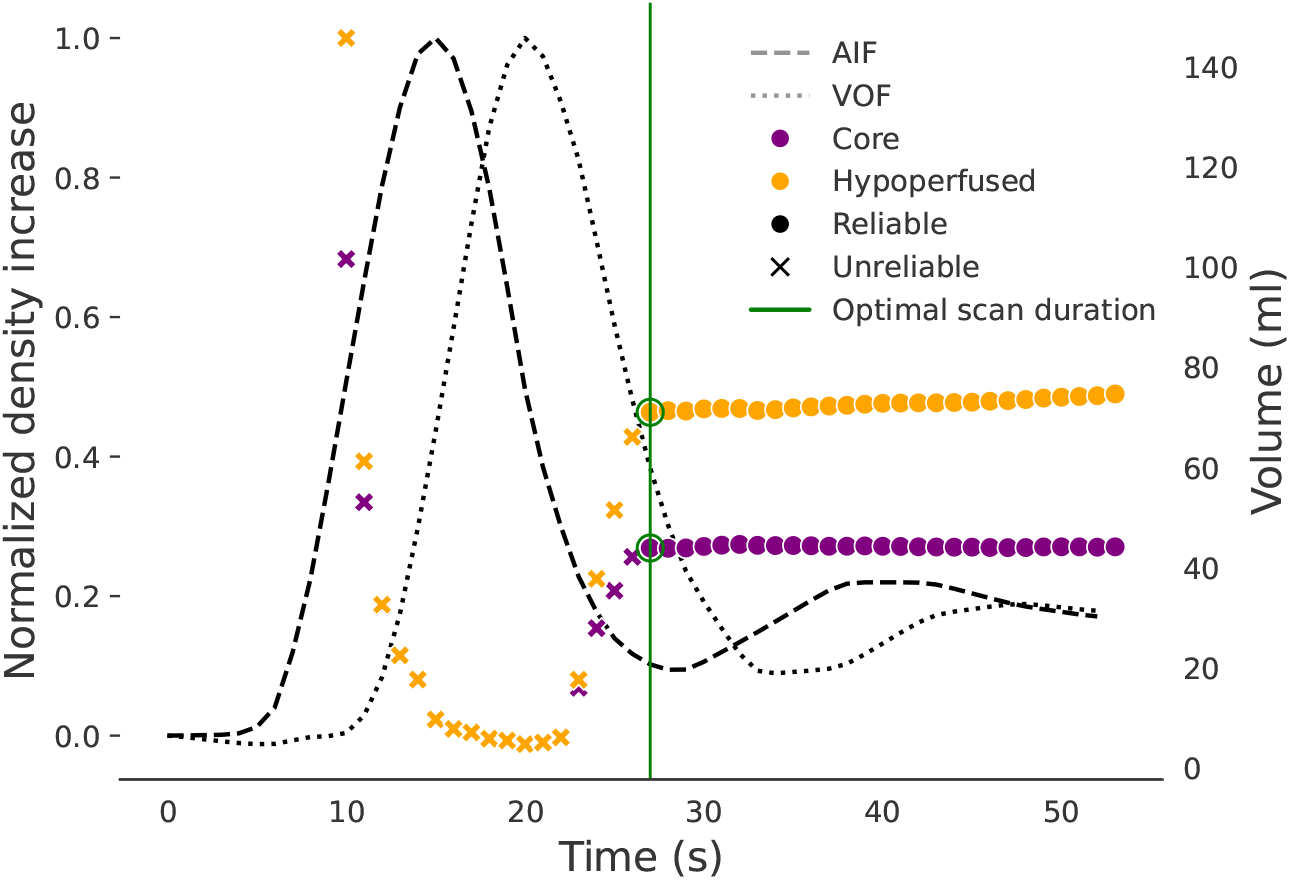
Reliable/unreliable lesion volumes computed at various scan durations. The arterial input function (AIF) and the vascular output function (VOF) are displayed as reference.

### Machine Learning for CTP Truncation Detection

Machine learning algorithms have been widely used to assess the quality of medical images Menze et al. (2008); Kyathanahally et al. (2018); Wei et al. (2019). We explore different machine learning models that could detect unreliably truncated scans by solely using information extracted from the vascular functions. The benefits of using the AIF and VOF to detect truncation artifacts are two-fold. First, the perfusion curves are always available in this imaging modality. Second, as they cover the entire perfusion event (note that these curves represent the contrast concentration inlet and outlet to the brain), they contain rich information for the problem under study. Consequently, it is needed to extract meaningful perfusion features that are impacted by an insufficient scan duration and that are, also, predictive of the truncation artifacts. Those features should capture the perfusion phases of the contrast-agent wash-in and wash-out and, ideally, they should be unaltered by the different CTP protocols used in clinical routine.

### Feature Extraction

All the explored machine learning algorithms are fed with the following 9 AIF/VOF derived features:

- Scan duration
- AIF/VOF time to the peak of the function (argmax*{*AIF*}*, argmax*{*VOF*}*)
- The *AIF/VOF coverage*, defined as the time difference between the peak of a signal and the scan duration:
  ∗ AIF_coverage_ = scan duration - argmax*{*AIF*}*
  ∗ VOF_coverage_ = scan duration - argmax*{*VOF*}*
- AIF/VOF upward and downward contrast increase
  ∗ AIF_UCI_ = AIF_t=argmax{AIF}_ - AIF_t=0_
  ∗ AIF_DCI_ = AIF_t=argmax{AIF}_ - AIF_t=scan duration_
  ∗ VOF_UCI_ = VOF_t=argmax{VOF}_ - VOF_t=0_
  ∗ VOF_DCI_ = VOF_t=argmax{VOF}_ - VOF_t=scan duration_

All features are visually represented in Fig 2.

**Figure 2:**
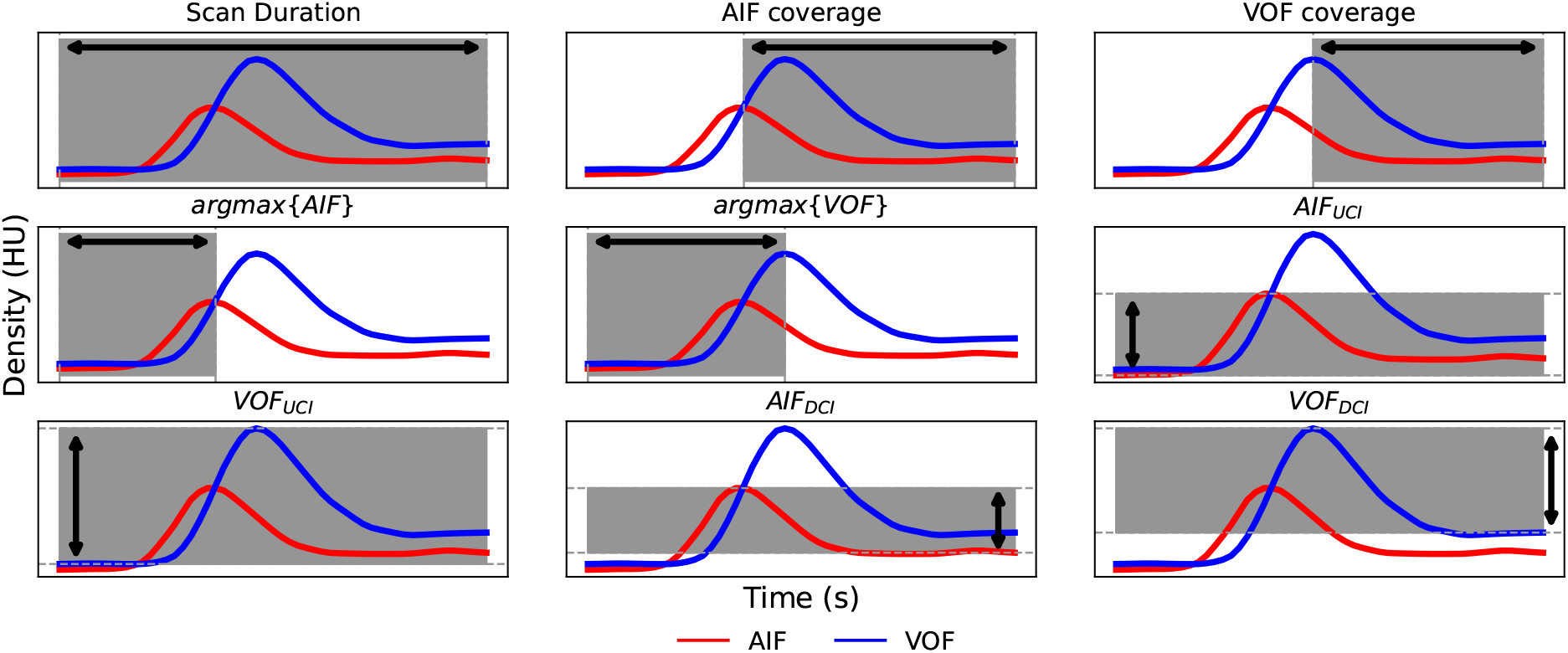
AIF and VOF derived features used to feed the machine learning algorithms. AIF: arterial input function; VOF: venous output function; HU: Hounsfield units; UCI: upward contrast increase; DCI: downward contrast increase.

### Classifiers & Model Fitting

We train six statistical/machine learning classifiers with the aim of detecting *reliable* and *unreliable* truncated scans. The trained models make use of linear or non-linear decision functions and are: *i*) random forests, *ii*) multivariate logistic-regression, *iii*) support vector machines with linear kernel, *iv*) support vector machines with radial basis kernel, *v*) Adaptive boosting (aka, Adaboost Freund and Schapire (1997)) and *vi*) Gradient boosting Friedman (2001). All models are trained using the scikit-learn python library Pedregosa et al. (2011).

### Data Augmentation

We augment our training dataset by generating synthetic samples in order to: *i*) compensate for the class imbalance between *reliable* and *unreliable* truncation samples and *ii*) model variable pre-contrast agent duration and variable contrast increases of the perfusion curves. Note that a different timing in the contrast bolus arrival alters the CTP scan duration but does not alter the presence of truncation artifacts. Likewise, the AIF and VOF contrast increase depends on the contrast agent iodine concentration. However, as the deconvolution algorithm is independent from the AIF/VOF absolute amplitudes, a variable vascular contrast increase does not alter the presence of truncation artifacts.

Balanced-class training sets are obtained using K-means SMOTE Last et al. (2017), a variation of the original synthetic minority oversampling technique Chawla et al. (2002), using the implementation in the Imbalanced-learn python library Lemaître et al. (2017). Simulation of contrast injection protocol variations is conducted by perfusion-specific data augmentation as similarly done in Robben and Suetens (2018); de la Rosa et al. (2021). Uniform distributions are used to randomly modify the pre-contrast agent duration and vascular contrast increases. When simulating variable pre-contrast duration, pre-contrast timing dependent features are increased or decreased by the same random factor (argmax{AIF}, argmax{VOF} and *scan duration*). For modelling variable contrast increases, the features AIF_UCI_, AIF_DCI_, VOF_UCI_ and VOF_DCI_ are scaled by a random factor.

### Experiments

We perform a 5-fold cross-validation experiment using an 80-20% train-test data split. The data splitting is conducted at the *scans* level, assuring that *i*) all untruncated and truncated versions of a same scan belong to the same fold and *ii*) the same data-splits are used to fit all the considered models. Only the training data is used parametrise the models and to select the classifiers’ operating point. Truncation predictions are later inferred over the unseen test data.

Besides, we compare the machine learning models against a baseline classifier which solely uses the *scan duration* as discriminant-rule. The classifier *g* operates as follows:

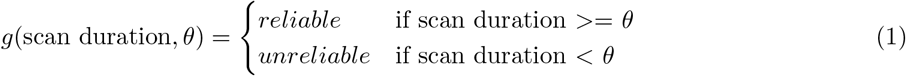

with *θ* a scan duration cutoff. This baseline is motivated by the CTP guidelines, which only consider the duration of a scan to avoid truncation artifacts Christensen and Lansberg (2019). Specifically, these guidelines suggest a cutoff of *θ* = 60 seconds in Equation 1. For our experiments, we evaluate the baseline classifiers’ performance at *θ* = [30, 40, 50, 60] *s*.

In order to understand the relevance of the AIF-VOF extracted features to discriminate *reliably* and *unreliably* truncated scans, we conduct a bootstrapping experiment by resampling 1,000 times the original database. In each iteration, a sample was drawn with replacement and was used to fit a classifier as described in Section *Classifiers & Model Fitting*. The relative feature importance is measured as defined in Friedman (2001) for decision tree ensembles. Briefly, the feature importance is calculated at the classifiers’ tree level as the impurity decay across all the nodes where that feature was used to create a split Kazemitabar et al. (2017). The final feature importance is computed as the average feature importance over all the considered trees. The mean and standard deviation feature importance for all features are reported. The chosen classifier for this experiment is the best performing one in terms of precision-recall AUC.

### Performance evaluation

The mean, standard deviation, 5th-95th percentiles and minimum and maximum of the scan duration and the optimal scan duration are reported for the entire dataset. The different algorithms’ performance are evaluated by conducting receiver-operating-characteristic (ROC) and precision-recall (PR) analysis. The area under the ROC and precision-recall curves are used as general classifier performance metrics. Besides, we measure the binary classification performance at the operating point closest to an ideal classifier with *precision = recall = 1*. The operating point is chosen from the fitted classifier as 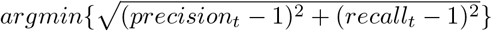, with *t* different classifier thresholds. Performance is measured in terms of precision 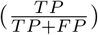, recall 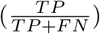 and F1-score 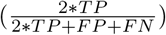, where acronyms represent TP: true positives, TN: true negatives, FP: false positives and FN: false negatives. The same binary classification metrics are reported for our baseline scan duration classifier, by making use of cutoffs *θ* = [30, 40, 50, 60] *s*. For these defined metrics, an *unreliable* truncation sample is considered as positive and a *reliable* truncation sample as negative.

## Results & Discussion

From the 156 analyzed scans, 132 scans (84.6%) are retained for further analysis. The remaining scans are discarded as 18 (11.5%) are unstable and 6 (3.9%) are free from CTP lesions. A total of 4954 synthetically truncated scans are obtained from the retained stable cases, from which 2640 (53.3%) are labelled as *reliable* and 2314 (46.7%) as *unreliable* (imbalance ratio of *∼*1.15:1, *reliable*:*unreliable*).

Descriptive statistics about the optimal scan duration are summarized in Table 1. It can be appreciated that a *∼*40-second scan duration suffices to get accurate perfusion volumes in 95% of our database, representing a much shorter acquisition than the 60-second guidelines recommendations Christensen and Lansberg (2019). These results differ from the reported ones in literature. Results obtained by Kasasbeh et al. (2016) and later adopted in the CTP guidelines Christensen and Lansberg (2019) suggest a 60 second scan duration to get reliable perfusion volumes in 90% of their cases. Copen et al. (2015) found severe truncation artifacts over Tmax when reducing perfusion MR acquisitions up to a 40-second scan duration. Their analyses identified Tmax (CBF) lesion reversal – defined by the authors as the false creation of a lesion on healthy areas or vice versa – in at least 42% (2%) of the shortened scans. Likewise, Borst et al. (2015) found truncated curves in 48-second duration scans (*∼*67% of scans with truncated AIF/VOF and *∼*20% of scans with truncated time attenuation curves in the core area).

**Table 1:** Descriptive statistics of the CTP scans. SD: Scan duration; OSD: Optimal scan duration. Std: standard deviation; P: percentile. AIF: arterial input function. All metrics are reported in seconds.

| | SD | OSD | (OSD - $\text{argmax}\{\text{AIF}\}$) |
| --- | --- | --- | --- |
| Mean (std) | 46.5 (5.2) | 27.5 (6.53) | 19.3 (7.22) |
| (Min, Max) | (31, 64) | (11, 43) | (2, 38) |
| (P5<sup>th</sup>, P95<sup>th</sup>) | (43.0, 56.6) | (18.1, 38.9) | (9.1, 32.0) |

The found variability on the minimal suggested scan duration across studies might come from different sources, namely: *i*) the type of deconvolution used in the experiments, *ii*) the biological and physiological variability of the patients (e.g. patient size and the cardiac output alters the contrast delivery through the brain Copen et al. (2015)), *iii*) physiopathological conditions that prolong the contrast-agent passage through the affected tissue, as happening in the hypoperfused tissue due to the ischemic occlusion Mikkelsen et al. (2015); Campbell et al. (2011) or in patients with severe intracranial vascular narrowing or multiple intracranial emboli Mangla et al. (2014), and *iv*) the contrast injection and CTP acquisition protocols (e.g. contrast injection rate, the pre-contrast scanning duration, syncronization between contrast injection and acquisition, etc.). Given all these sources of variability, using a fixed minimal scan duration might sometimes be enough to accurately measure perfusion volumes though it might truncate CTP scans in need of longer acquisitions for any of the previously listed reasons.

It is worth to mention that the optimal scan duration reported in Table 1 is also dependent on the pre-contrast duration. Since pre-contrast duration is not standardized in clinical practice, we compute the time difference between the optimal scan duration and the AIF peak (argmax {AIF}), as it is more informative than the optimal scan duration and less biased by the different CTP protocols. In Table 1 this metric is reported for our entire database. Results show that 95% of our scans require 32 seconds following the AIF time-peak to obtain reliable perfusion volumes. Please note that this metric is pre-contrast protocol independent but still influenced by several other variables (as the deconvolution software and the patients’ characteristics).

### Truncation Artifacts Detection

Figure 3 shows the ROC and precision-recall curves obtained with the different classifiers when differentiating *reliable* from *unreliable* truncated acquisitions. Overall it can be seen that classifiers yielded a similar high performance for both the considered metrics. The gradient boosting slightly outperformed the remaining classifiers with an ROC-AUC of 0.964 and a precision-recall AUC of 0.959. It is also worth pointing out that the multivariate logistic regression classifier obtained a lower performance than the baseline classifier, which solely uses the *scan duration* as input feature.

**Figure 3:**
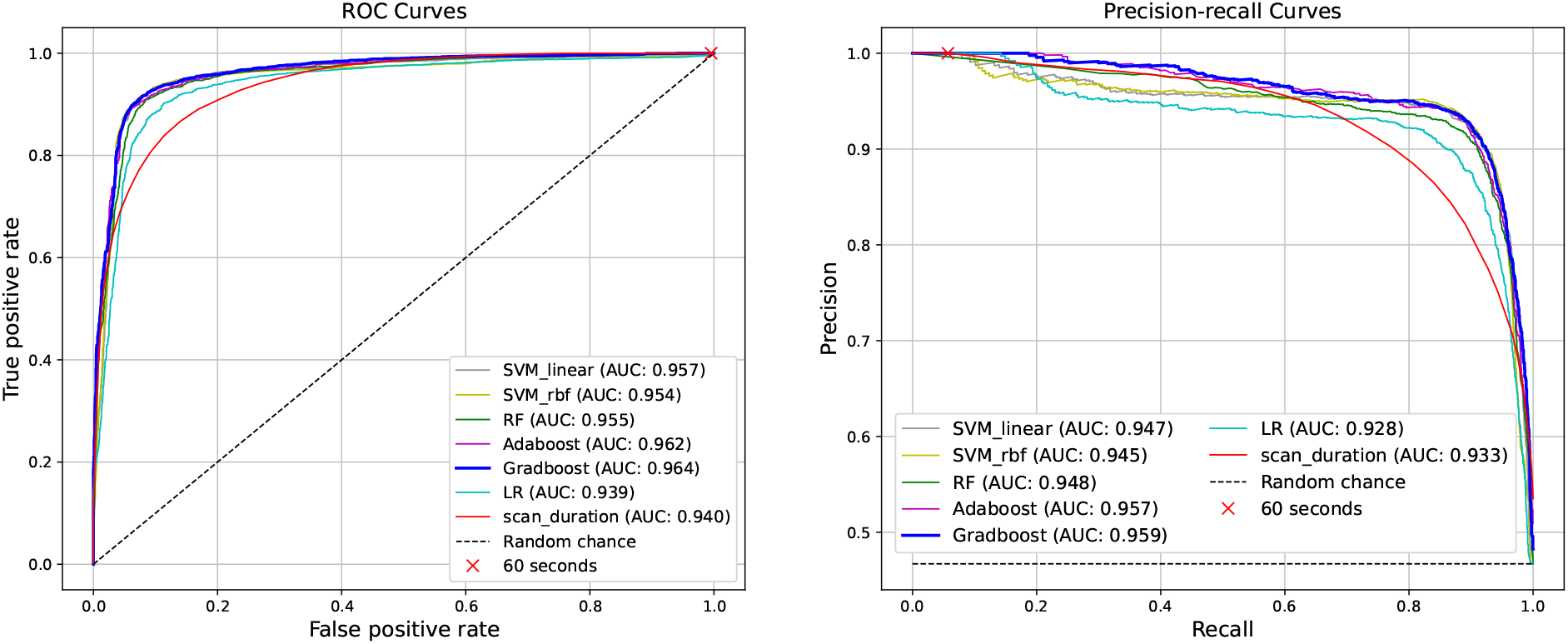
Receiver operating characteristic (left) and precision-recall (right) curves. AUC: area under the curve; SVM_linear: support-vector machine with linear kernel; SVM_rbf: support-vector machine with radial basis function kernel; RF: Random forests; LR: Logistic-regression; Adaboost: Adaptive boosting; Gradboost: Gradient boosting.

When assessing the classifiers capability for detecting truncation artifacts at the chosen operating point, the results of Table 2 are obtained. Machine learning models yielded very similar performance for the considered metrics and have considerably outperformed the baseline classifier *g*(scan duration, *θ* = 60s). It is also worth noting that models with lower ROC and precision-recall AUCs (such as SVM_linear_ and SVM_rbf_, Fig. 3) have provided slightly better results at the operating point compared to the Gradient boosting classifier. The highest F1-score for detecting unreliable perfusion volumes is achieved with a support vector machine with radial-basis-function kernel (F1-Score=0.913). In Fig. 4 we illustrate, for the outperforming classifier in terms of ROC and precision-recall AUC, the distribution of the predicted samples in terms of their scan duration to optimal scan duration difference (Fig. 4). It can be appreciated that all the mis-classifications are bounded in an approximate interval of *±* 15 s. Thus, within this temporal window the classifier struggled the most to correctly detect unreliable perfusion volumes and, outside this temporal window, the classifier correctly predicted all samples. Besides, as expected, the closer a scan duration is to its optimal scan duration (i.e, the scan duration approximates to the inflexion point where *unreliable* samples become *reliable*), the harder for the model is to correctly classify a sample. Nonetheless, the absolute frequency of incorrect classifications is always much smaller than the absolute frequency of correct ones, independently of a scan duration’s closeness to its optimal scan duration.

**Table 2:** Classifiers’ performance for detecting truncation artifacts. The used operating points are *θ* = [30, 40, 50, 60] *s* for the baseline classifier using the *scan duration* information. For the machine learning approaches, the chosen operating point is the one closest to the ideal classifier with *precision = recall = 1*. Outperforming values for each metric are shown in bold. SVM_linear: support-vector machine with linear kernel; SVM_rbf: support-vector machine with radial basis function kernel; Gradboost: Gradient boosting classifier.

| Classifier | Recall | Precision | F1-Score |
| --- | --- | --- | --- |
| SVM<sub>linear</sub> | 0.908 | 0.913 | 0.910 |
| SVM<sub>rbf</sub> | 0.906 | 0.921 | **0.913** |
| Random Forests | 0.888 | 0.918 | 0.903 |
| Adaboost | 0.901 | 0.914 | 0.908 |
| Gradboost | 0.903 | **0.922** | 0.912 |
| Logistic Regression | 0.897 | 0.877 | 0.887 |
| scan duration $\theta=30s$ | 0.928 | 0.775 | 0.844 |
| scan duration $\theta=40s$ | 0.997 | 0.568 | 0.724 |
| scan duration $\theta=50s$ | **1.000** | 0.478 | 0.647 |
| scan duration $\theta=60s$ | **1.000** | 0.469 | 0.638 |

**Figure 4:**
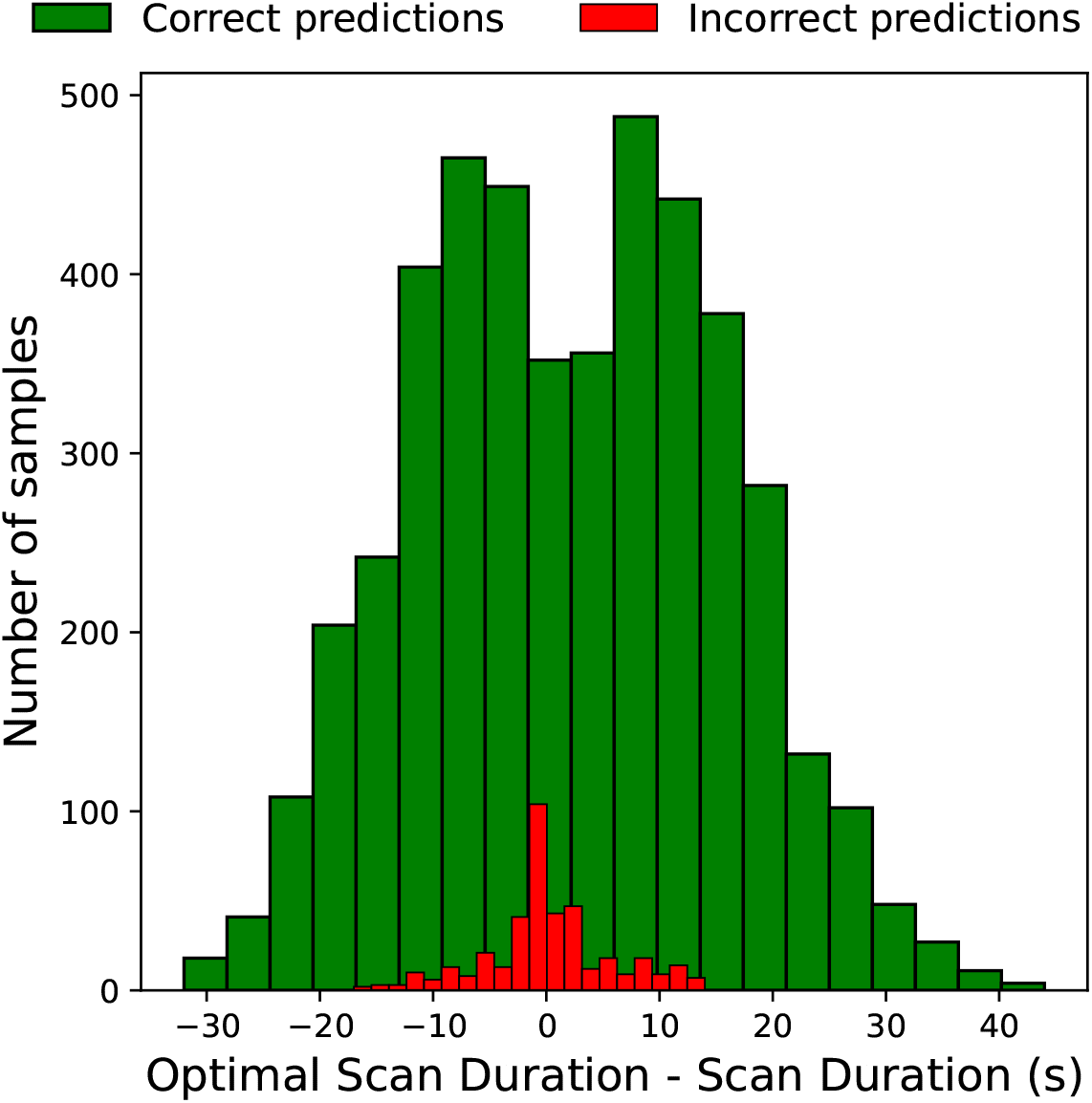
Histogram showing the difference between optimal scan duration and the scan duration for each predicted sample using a Gradient boosting classifier. Samples are grouped by their prediction status (correct/incorrect). Correctly predicted samples comprise true positives and true negatives. Incorrectly predicted samples comprise false positives and false negatives.

For the baseline classifier *g*, the highest detection performance is obtained at a cutoff *θ* =30 s (F1-score = 0.844). When using the clinical standard cutoff *θ*=60 s, the baseline classifier showed the maximal recall of 100% for detecting unreliably truncated scans. These results are expected as the optimal scan duration for the ISLES’18 dataset have much lower values than 60 s (Table 1). Although *g*(scan duration, *θ*=60 s) is extremely efficient at detecting truncation artifacts, it does it at the expense of generating many false positives (low precision of 0.469 and a low F1-Score of 0.638, Table 2). The ROC and precision-recall operating points at *θ*=60 s are shown in Figure 3. It can be appreciated that this operating point falls on the boundaries of the classifiers’ ROC and precision-recall curves. On one hand, a 60s scan duration could be a safe recommendation under fixed acquisition and post-processing considerations (as deconvolution type, injection and acquisition protocol) to avoid truncation artifacts, though it could still expose patients to unnecessary ionizing radiation when these considerations do not hold. On the other hand, when performing quality analysis on already acquired CTP scans, the 60 s scan duration is a very poor criterion to identify truncation artifacts as it does not consider any of the truncation confounders listed above. Our experiments show that machine learning models fed with perfusion-derived features can unveil acquisitions affected by truncation and reliably detect erroneous perfusion measurements. The proposed features are simple, robust to extract even in low quality acquisitions and independent (except for the *scan duration*) from the contrast injection and CTP acquisition protocols.

### Importance of the AIF and VOF Features

Figure 5 summarizes the different features’ relevance obtained when fitting 1,000 Gradient boosting classifiers in a resampling with replacement bootstrapping fashion. The AIF_coverage_ shows to be the most crucial feature for detecting *unreliable* perfusion volumes due to truncated acquisitions. Besides, the VOF_DCI_, VOF_coverage_ and AIF_UCI_ also result to be important features for the machine learning model. The large predictive value of the AIF_coverage_ and the VOF_coverage_ features can be related to their robustness to variable pre-contrast agent durations. The *scan duration* feature, instead, is affected by the CTP acquisition protocols and as such, shows less relevance for the fitted models. The contrast increase features AIF_UCI_ and VOF_DCI_ are useful for the task as they represent the beginning and ending of the agent delivery through the tissue, allowing the classifier to capture the characteristics of an entire or truncated perfusion event.

**Figure 5:**
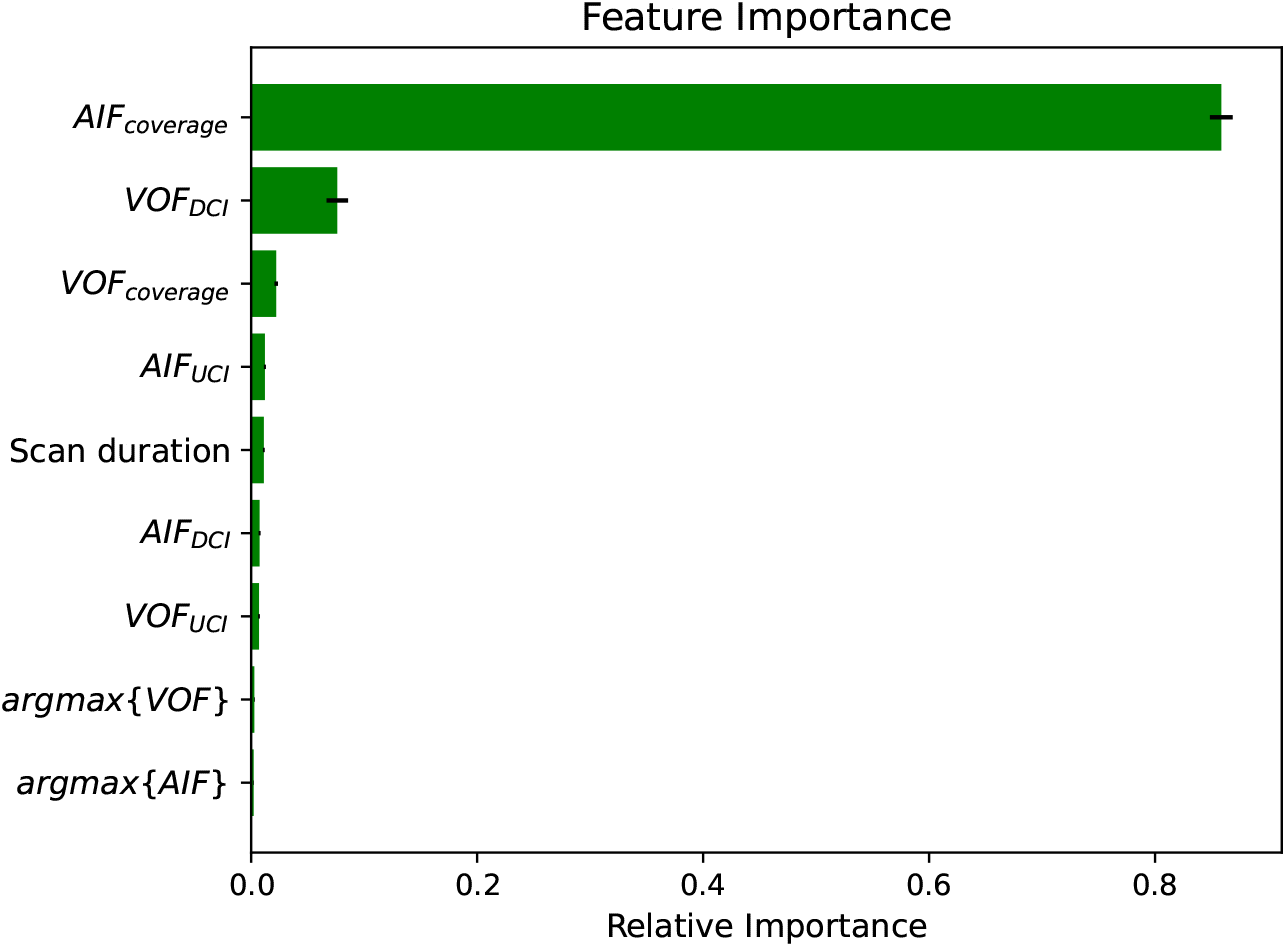
Relative feature importance for 1,000 bootstraps with a Gradient boosting classifier. Bars (error-bars) represent mean (standard deviation). AIF: arterial input function; VOF: venous output function; UCI: upward contrast increase; DCI: downward contrast increase.

Based on these results we finally explore whether the most relevant feature, AIF_coverage_, might have discriminant power to detect truncation artifacts. To this end, AIF_coverage_ is used instead of the *scan duration* to build a new classifier *g*^*′*^(AIF_coverage_, *ρ*) that operates as described in Eq. 1. While the *scan duration* based classifier *g* obtained a ROC and precision-recall AUCs of 0.940 and 0.933 respectively, the classifier *g*^*′*^ using AIF_coverage_ as unique feature yielded 0.960 and 0.949 ROC and precision-recall AUCs respectively. These results evidence that AIF_coverage_ is a strong discriminant feature for detecting truncation artifacts. Besides, the close performance of *g*^*′*^ to the results obtained with machine learning models (Fig. 3) show that these algorithms mostly predict outputs based on the AIF_coverage_ information and get some extra benefit from the remaining perfusion features.

### Effect of the Data Augmentation

Finally we conduct an ablation study to assess the effect of the two used data augmentation approaches to train the machine learning models. For this experiment, a 5-fold cross validation scheme is followed by training a Gradient boosting classifier in three ways: *i*) With the original, un-augmented dataset, *ii*) Applying perfusion-specific data augmentation by modelling variable contrast bolus arrivals and variable AIF/VOF contrast increases and *iii*) By applying perfusion-specific data augmentation and class-balancing with K-means SMOTE Last et al. (2017). Results are summarized in Table 3. It can be seen that both the ROC and the precision-recall AUC improve when simultaneously using both types of data-augmentation to generate synthetic samples.

**Table 3:** Data augmentation effect over the Gradient boosting classifier performance. DA: Data Augmentation. K-SMOTE: K-means variant of the Synthethic Minority Oversampling Technique Last et al. (2017); ROC: receiver operating characteristic. PR: precision-recall; AUC: area under the curve.

|  | No DA | Perfusion DA | Perfusion DA + K-SMOTE |
| --- | --- | --- | --- |
| ROC-AUC | 0.961 | 0.963 | **0.964** |
| PR-AUC | 0.954 | 0.958 | **0.959** |

### Limitations and Future Directions

There are some considerations about this research that should be cautiously taken. It is worth to mention that our conclusions only hold for CTP analysis using time-invariant singular value decomposition deconvolution. Other techniques used for perfusion analysis might behave differently under truncation scenarios. Still, the delay-invariant singular value decomposition deconvolution is the most widespread and used algorithm in software packages Fieselmann et al. (2011); Kudo et al. (2010); Vagal et al. (2019). Readers interested in the effect of CTP truncation over different parameter map estimation methods are referred to the work of Copen et al. (2015), as such inter-algorithm comparisons are out of the scope of this research. It is worth saying that while the devised models only hold for the ISLES’18 database characteristics and for the deconvolution algorithm used in this study, the extracted features are generalizable and allow the adaptation of these models to other deconvolution algorithms or imaging modalities (as perfusion MRI). In addition, the deployment of a truncation artifacts detection method in automatic CTP evaluation software is limited to the AIF/VOF selection performance. In this work, all the experiments have been conducted using manually annotated vascular functions. As such, failures in the CTP curves selection could produce a misleading truncation analysis using our proposed methodology. Nonetheless, recent approaches using dedicated artificial intelligence methods show efficacy and robustness to select vascular functions even under low quality CTP scenarios Winder et al. (2020); de la Rosa et al. (2021). Finally, future directions for this work might involve the machine-learning prediction of missing CTP time-points at the end of the series. As such, reconstructing the ending perfusion phase of the vascular functions could help improve the detection of truncation artifacts.

## Conclusion

We show that a *∼*40-second scan duration is sufficient to avoid truncation artifacts in 95% of the multi-center/scanner ISLES’18 dataset. However, solely using the *scan duration* criterion as a truncation artifacts avoidance is suboptimal. Depending on the patients’ physiology, the contrast injection and/or the CTP acquisition protocols, much shorter scan durations can still avoid truncation artifacts while scans with 60-70s duration can still lead to unreliable lesion volumes. To overcome this variability present in clinical routine, we have extracted and identified AIF and VOF derived features that are predictive of truncation artifacts. These features were shown to be fully (or at least more) independent from the centers’ acquisition protocols than the *scan duration*. Furthermore, machine learning models fed with the perfusion features yielded high performance for detecting unreliable lesion volumes due to truncation effects. We conclude that these methods could be transferred to CTP post-processing software and, as such, may increase the interpretability of CTP outputs in acute stroke settings.

## Data Availability

Data produced in the present study is not available.

## Acknowledgement

This project received funding from the European Union’s Horizon 2020 research and innovation program under the Marie Sklodowska-Curie grant agreement TRABIT No 765148.

